# Developing an open-source framework for LLM evaluation of patients using EHR clinical documentation; performance of LLMs relative to medical professionals

**DOI:** 10.64898/2026.08.21.26361031

**Authors:** Liam Barrett, Nikhil Joshi, Alexander S. North, Lilia Dimitrov, Elizabeth F. Maughan, Talisa Ross, Rahul Pankhania, Kiara Paramjothy, Iona Minty, Luiza Farache-Trajano, Sabrina L. Smith, Katrina A. Mason, Eishaan K. Bhargava, Catherine Donnelly, Hanaa Fatoum, Ashwika Padiyar, Zahra Kader, Chun Hin Kevin Chan, Anne GM Schilder, Nishchay Mehta

## Abstract

**Background:** Large language models (LLMs) have shown increasing capability in medical knowledge tasks, yet how they perform in extracting structured clinical information from real-world clinical documentation remains uncertain. We evaluated the performance of LLMs relative to medical professionals in extracting SNOMED-coded clinical information from openly available Ear, Nose and Throat (ENT) EHRs from MTSamples, examining both reliability and accuracy metrics.

**Methods:** We evaluated the performance of seven LLMs (including GPT-4o, Claude 3.5, Gemini 1.5 Pro, Gemma 3 and three LLAMA variants) against annotations from fourteen medical professionals who served as both study authors and data annotators. Each annotator independently extracted seven categories of clinical information from 98 publicly available ENT clinical documents: socio-demographics, symptoms, signs, diagnoses, treatments, risk factors, and test results. Standardised medical terminology was enforced through SNOMED-CT code assignment, enabling standardised comparison through Cohen’s Kappa. We employed Bayesian hierarchical modelling to test non-inferiority of medic-LLM agreement compared to medic-medic agreement, using Beta distributed likelihood functions with weakly informative priors. Non-inferiority margins of 0.05, 0.10, and 0.15 were assessed with 95% posterior probability thresholds.

**Results:** Cohen’s Kappa for inter-rater reliability was 0.752 (95% CI: 0.710 – 0.794) between medical professionals and 0.391 (95% CI: 0.362–0.420) between LLMs and medical professionals. Bayesian analysis showed medic-medic agreement (posterior mean 0.813, 95% CI: 0.755–0.860) exceeded medic-LLM agreement (0.659, 95% CI: 0.633–0.684) by 0.154 (95% CI: 0.091–0.209). Non-inferiority was rejected at all tested margins (*δ* = 0.05, 0.10, 0.15). Agreement varied by clinical category, with smallest differences for test results and largest for diagnoses. GPT-4o achieved 97.0% precision and 84.9% recall, with a 7.5% false positive rate.

**Conclusions:** Current LLMs do not achieve inter-rater reliability levels comparable to medical professionals in clinical information extraction from ENT documentation. These findings provide evidence-based guidance for LLM deployment in clinical documentation workflows, suggesting they are best suited for initial extraction with human verification rather than autonomous operation.

## Introduction

The digitisation of healthcare systems has produced a wealth of nuanced clinical documentation residing in electronic health records (EHRs) [1]. These digital clinical documents contain rich descriptions of patient symptoms, clinical findings, diagnostic considerations, and treatment decisions that are crucial for understanding disease presentation and progression, and treatment outcomes [2].

Extracting and processing structured and standardised data from the narratives in clinical documents at scale remains a significant challenge for both medical professionals. The complexity of medical language, domain-specific knowledge requirements and the idiosyncratic nature of how medical professionals generate EHRs [3] poses barriers for automated information extraction. Whilst it is already possible to use large language models (LLMs) on EHRs to prognosticate [4], predict outcomes [5, 6] and treatment responses [7], it is unclear whether the models attend to the same relevant information as medical professionals. Transformer-based LLMs have shown promising results on standardised clinical knowledge assessments such as the United States Medical Licensing Examination style questions (MedQA), with some models achieving performance comparable to medical professionals on licensing examinations [8]. For example, Med-PaLM performed with 67.6% accuracy on the MedQA, though important gaps remained in comparing model outputs to expert clinician annotations [9]. While these approaches have shown successes [9], their ability to extract clinically relevant information from real-world ENT, hearing and balance documentation remains relatively unexplored [10].

The assessment of LLM performance in clinical information extraction presents unique methodological challenges [11]. While traditional metrics like accuracy provide useful benchmarks, they may not fully capture the nuanced requirements of clinical documentation [12]. Inter-rater reliability measures, commonly used to evaluate consistency among medical professionals, offer a promising framework for assessing LLM performance against clinical ground truth. Furthermore, the use of standardised medical terminologies like SNOMED-CT (Systematised Nomenclature of Medicine – Clinical Terms) provides an objective standard for evaluation [13]. SNOMED-CT’s hierarchical structure and rich semantic relationships allow precise encoding of clinical concepts, from high-level disease categories to specific symptoms and procedures, enabling reproducible evaluation across different healthcare settings [8, 9].

This study is part of a broader research programme leveraging artificial intelligence to better understand the deep phenotype of ENT, hearing and balance conditions [14]. ENT, hearing health and balance conditions present an ideal test case for clinical AI evaluation due to its high patient throughput [15], complex terminology, and notably, its under- utilisation of deep phenotyping approaches [16] that are essential for emerging precision therapeutics in hearing loss [14, 16], balance disorders [17], and head and neck cancers [18, 19]. Our group has previously demonstrated the potential of machine learning approaches in this domain through systematic reviews of unsupervised phenotyping methods [16] and the creation of comprehensive health informatics resources for hearing research [20]. We first demonstrated the feasibility of using automated information extraction in ENT with simpler neural networks [21] however, further advances in performance are required for clinical use.

We present a systematic evaluation of LLM performance against medical professionals in extracting and processing clinical information from 98 ENT EHRs. Fourteen medical professionals and seven LLMs (including GPT-4o, Claude 3.5, Gemini 1.5 Pro, Gemma 3 and three LLAMA variants) independently extracted seven categories of clinical information: socio-demographics, signs, symptoms, diagnoses, treatments, risk factors, and test results. Standardised medical terminology was enforced through SNOMED-CT code assignment, enabling rigorous comparison through established inter-rater reliability metrics. We employed Bayesian hierarchical modelling to formally test whether LLM-medic agreement achieves non-inferiority compared to medic-medic agreement.

Through this investigation, we aim to advance the understanding of LLM capabilities in specialised clinical domains while establishing rigorous methodologies for evaluating their performance across healthcare applications [11].

## Methods

### Ethical Considerations

This study utilised publicly available, anonymised clinical documentation from MTSamples provided for research and training purposes. The clinical information extraction and annotation tasks were performed by the authors of this study who are medical professionals (N.J., A.N., L.D., E.M., T.R., R.P., K.P., I.M., L.F.T., S.S., K.M., E.B., C.D., H.F.), all of whom provided informed consent for their participation in the data annotation process. As the annotation work was conducted by study authors analysing publicly available, de-identified data, this research design is analogous to systematic review methodology where authors serve as data extractors.

Following consultation with the University College London Research Ethics Committee guidelines, it was determined that formal ethics approval was not required for this study design. While ethics approval was not required, to ensure appropriate data handling and processing practices, we registered our data processing protocols with University College London’s Data Protection Office who approved our data management procedures (Reference: Z6364106/2025/01/13). This registration ensures compliance with data protection regulations while confirming adherence to the principles of the Declaration of Helsinki for research involving human data.

### Study Design

We conducted a cross-sectional study to assess inter-rater reliability and validity of clinical information extraction of LLMs from ENT EHRs. The study employed a structured annotation framework to evaluate both the reliability and validity of automated information extraction against medic performance.

### Dataset

The dataset comprised of 98 EHRs sourced from the ENT section of MTSamples (https://mtsamples.com). These EHRs were pre-processed and standardised into a consistent JSON format, with each entry containing the EHR text, medical specialty classification, and associated metadata. For detailed preprocessing steps and data structure, see §Code availability.

### Annotators

#### Medical Professional Annotation

Fourteen authors of this study (N.J., A.N., L.D., E.M., T.R., R.P., K.P., I.M., L.F.T., S.S., K.M., E.B., C.D., H.F.), all qualified doctors with a minimum of 6 months ENT experience, performed independent annotations on the clinical documentation. Each annotator provided informed consent for their participation and was aware that their annotation data would be analysed and reported in aggregate form. The annotation process utilized an online testing platform that mirrors the LLM prompting protocol to ensure methodological consistency.

The 98 test documents were pseudo-randomly distributed among the annotators, with each document being reviewed by a minimum of two independent medical professionals. To minimise systemic pair bias across the dataset, documents were assigned using a modified Hungarian algorithm for bipartite matching optimisation [22], designed to maximise annotator pair diversity while ensuring an even workload distribution. All annotators completed their assessments independently without access to other annotators’ responses.

#### Large Language Models

We evaluated seven large LLMs (GPT-4o, Sonnet 3.5, Gemini 1.5, LLAMA 3.1 (405B; 70B; 8B) and Gemma) across the three above categories (State-of-the-Art (SOTA), Large Open-Source and, Local Deployment Models) to systematically assess the relationship between model architecture, scale, and clinical information extraction performance.

Three cloud-based proprietary models were evaluated via their respective APIs. GPT- 4o (OpenAI, version: gpt-4o-2024-08-06), A multimodal transformer model with enhanced reasoning capabilities, accessed via the OpenAI API. Maximum context length was set to 8,192 tokens. Claude 3.5 Sonnet (Anthropic, version: claude-3-5-sonnet-20241022), An advanced language model optimised for complex analytical tasks, accessed via the Anthropic API. Maximum output length was configured to 8,192 tokens. Gemini 1.5 Pro (Google, version: gemini-1.5-pro), A multimodal model with extended context capabilities, accessed via the Google Generative AI API.

Three open-source models from Meta’s Llama 3.1 family were evaluated via cloud deployment. Llama 3.1 405B (meta/llama-3.1-405b-instruct-maas) was the largest publicly available language model at the time of evaluation, with 405 billion parameters. Llama 3.1 70B (meta/llama-3.1-70b-instruct-maas), A mid-scale variant balancing computational efficiency with performance. Llama 3.1 8B (meta/llama-3.1-8b-instruct-maas), the smallest Llama variant tested, designed for resource-constrained deployments.

One model was evaluated for potential on-premises healthcare deployment: Gemma 3 12B (google/gemma-3-12b-it). A 12-billion parameter model designed for efficient local deployment.

To ensure reproducible and deterministic outputs across all models, we implemented strict parameter control:

- **Random Seed**: Fixed at 42 across all compatible APIs
- **Top-p sampling**: Disabled (effectively 1.0) to ensure deterministic token selection
- **Output tokens**: Set to 8,192 tokens to allow for standardised cross-comparisons.

All models were accessed programmatically via their respective REST APIs using authenticated HTTPS connections. API versions were fixed at the time of evaluation (Q1 2025) to ensure consistency. Rate limiting was implemented to respect API quotas, with automatic retry logic for transient failures. Response parsing included validation of JSON structure and SNOMED-CT code format compliance.

The consistent configuration across models enabled fair comparison while the range of model scales (8B to 405B parameters) and deployment modes (proprietary cloud, opensource cloud, local-capable) provided comprehensive coverage of current LLM capabilities for clinical applications.

Full specification and documentation of how these models were implemented are available at the paper’s codebase (see §Code availability).

### Procedures

#### Annotation Framework

The annotation process was designed to extract seven key categories of clinical information:

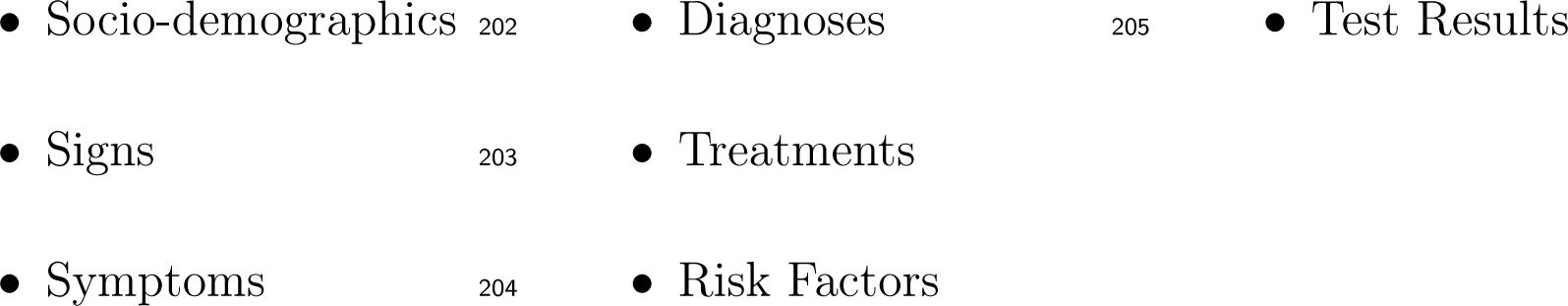

For each identified element, annotators provided mandatory fields including the exact text, surrounding context, and additional qualifiers such as laterality, presence status, and temporal information. Standardised medical terminology was enforced through SNOMED-CT code assignment (See Table 1 for example annotations). The SNOMED-CT UK database 39.0.0 was used.

**Table 1:**
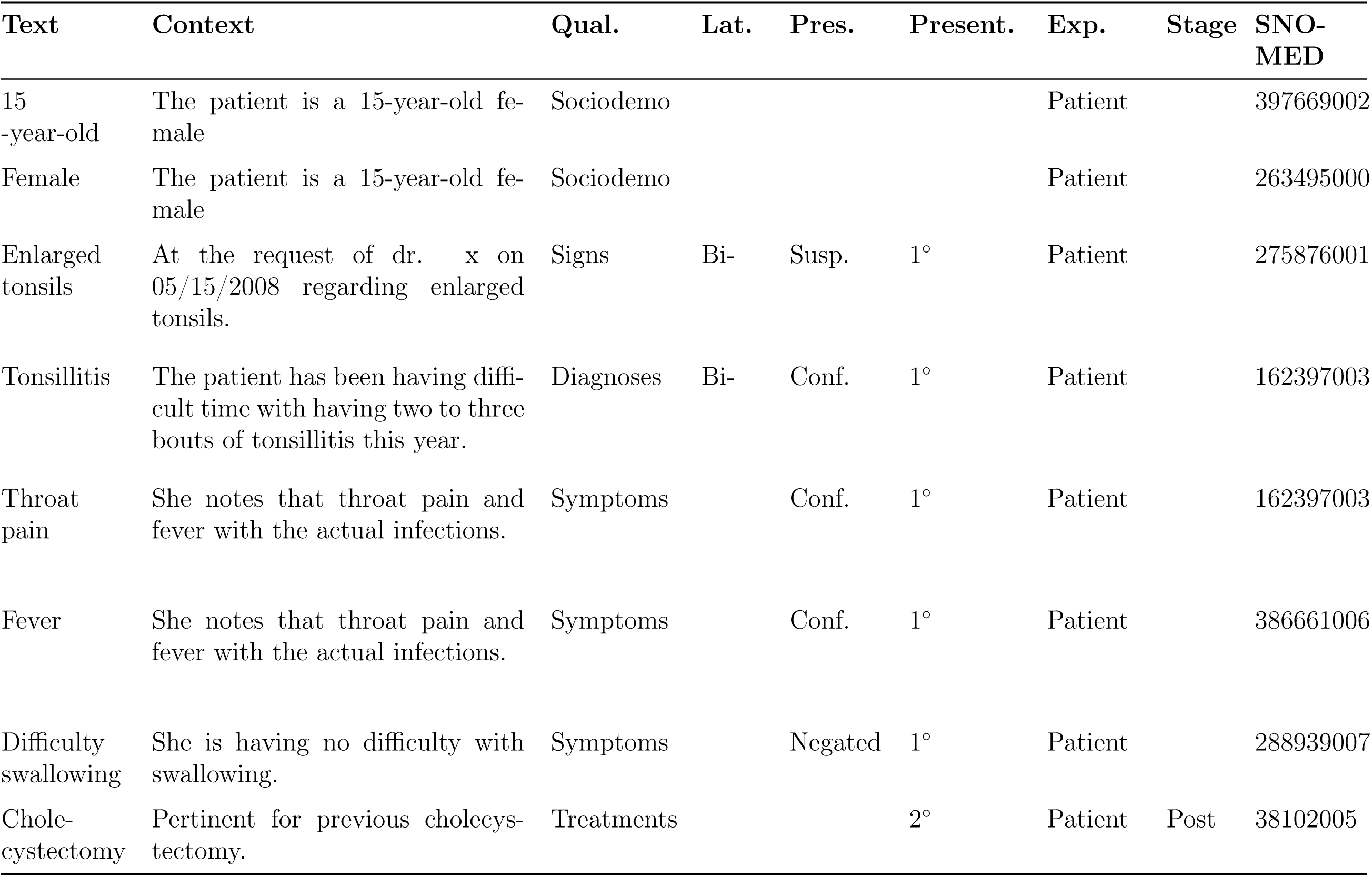
Example Labels, meta-data and qualifiers for EHR 0018.

#### LLM Annotation

We evaluated three categories of LLMs to assess the relationship between model scale and performance: State-of-the-Art (SOTA), Large Open-Source and, Local Deployment Models.

For SOTA models, we used cloud-based proprietary models including OpenAI’s GPT- 4o, Anthropic’s Sonnet Claude 3.5, and Google’s Gemini 1.5 Pro were tested with standardised parameters. The models and the chosen parameters are fully specified in section §Large Language Models.

Similar to the SOTA models we also tested a series of large open source models which allow the researchers more fine grained control while maintain the similar inferential power as proprietary, SOTA models. Cloud-deployed open-source models including Meta’s Llama 3.1 variants (405B and 70B parameters) were evaluated under identical conditions. Finally, we also assessed smaller, locally deployable models including Llama 3.1 (8B), and Gemma 3 (12B). These were tested to assess feasibility for closed healthcare deployments.

All models utilised a chain-of-thought prompting strategy with one-shot learning, implemented through a standardised five-step process (Figure 1).

**Figure 1:**
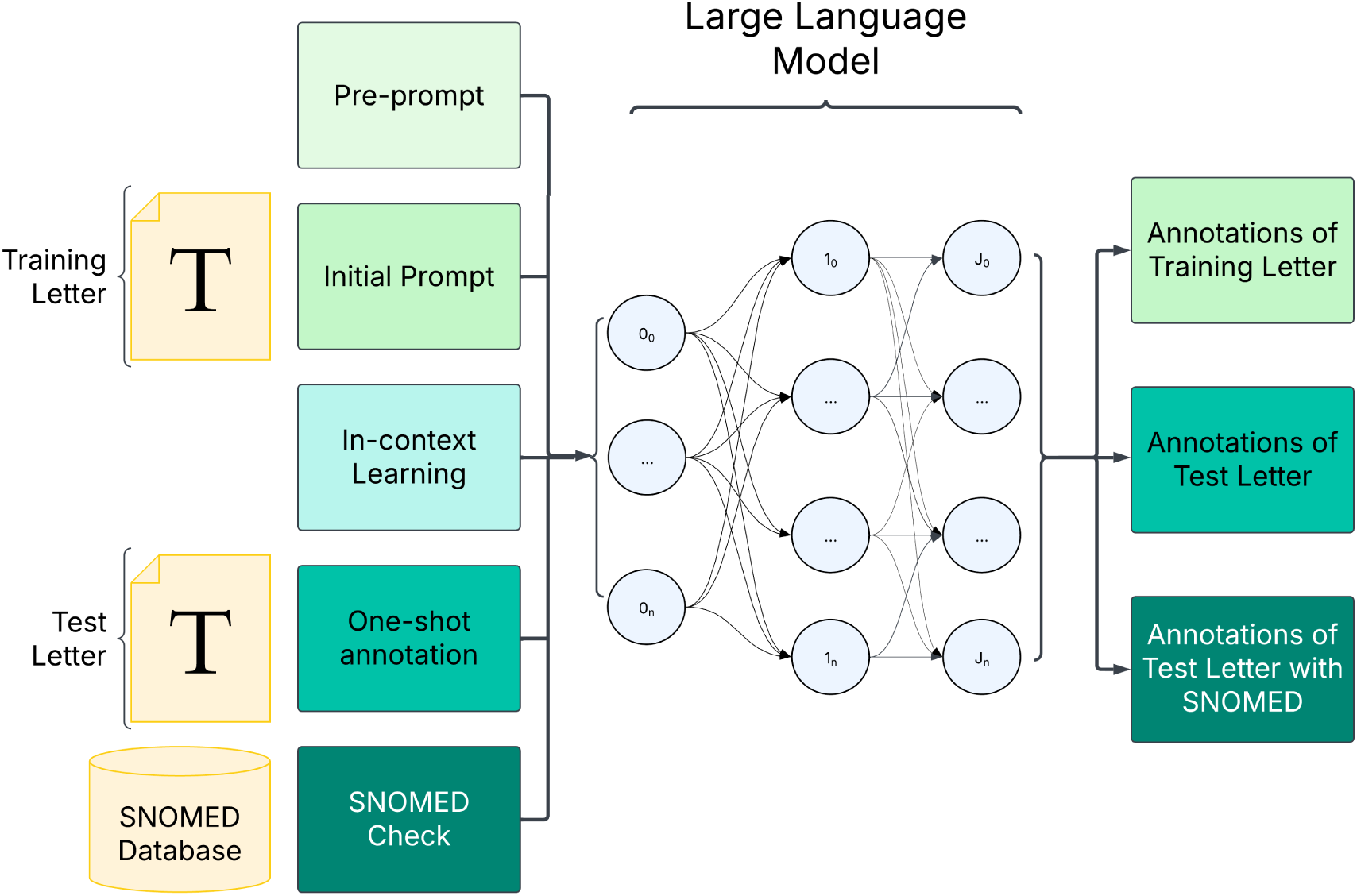
LLM Clinical EHR Annotation Process Flow. The process involves multiple steps: (1) Pre-prompting the model with medical expertise instructions, (2) Initial prompt with the task description, (3) In-context learning using a training EHR and its annotations, (4) One-shot annotation of the test EHR, and (5) SNOMED code validation using the SNOMED database

This compromised of:

1. **Pre-prompt:** The model is pre-prompted that they are an expert in medical and clinical fields. They are tasked with analysing EHRs and extracting relevant labels from these EHRs.
2. **Initial prompt:** They are prompted with the task (labelling EHRs).
3. **In-context learning:** They are prompted with an example EHR (EHR 0080, available in [23]) to annotate. The annotations are retrieved and stored. The LLM is then provided the full example annotation of EHR 0080 as feedback (available in §Code availability).
4. **One-shot annotation:** They are then prompted with the entire EHR and asked to generate a complete list of labels and key text associated with that label, as well as qualifiers and meta-data (See Table 1). This should yield the LLM’s first attempt at labelling the EHR.
5. **SNOMED-CT API call:** The key text is then put to SNOMED-CT API to retrieve the associated SNOMED code programmatically. The LLM is then provided with the results of the SNOMED-CT API call and is prompted to decide which code(s) are appropriate. If the LLM reports that the codes are inappropriate, the LLM can specify a new search term for the appropriate code once.

This procedure then provides the researcher with the outputted annotation

### Statistical Analysis

The objectives of this work are formalised in two primary hypotheses:

H1: There is no significant difference in inter-rater reliability between LLMs and medical professionals when extracting SNOMED-coded clinical information from ENT EHRs.

H2: There is no significant difference in inter-rater reliability between LLMs and medical professionals when extracting SNOMED-CT codes across different clinical information categories.

We employ Bayesian hierarchical modelling to evaluate our two primary hypotheses regarding LLM performance relative to medical professionals. For inter-rater reliability analysis, we treat each pairwise Cohen’s Kappa value as a sample from underlying group-specific distributions, allowing us to compare the distribution of medic-medic (M-M) agreement against medic-LLM (M-L) agreement.

For Hypothesis 1, we construct a hierarchical Bayesian model where pairwise Cohen’s Kappa values are modelled using a Beta distribution to respect the [0,1] bounds of the coefficient:

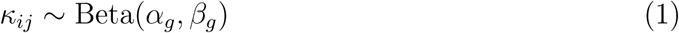

where *κ_ij_* represents the Cohen’s Kappa between annotators *i* and *j*, and *g ∈ {*M-M, M-L*}* indicates the comparison group. The shape parameters *α_g_* and *β_g_* are given weakly informative priors. Non-inferiority is assessed using a margin of *δ* = 0.1, with strong evidence for non-inferiority defined as *>* 95% posterior probability that the mean M-L agreement exceeds mean M-M agreement minus *δ*.

For Hypothesis 2, we extend the model to incorporate clinical categories:

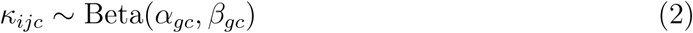

where *c* indexes the seven clinical categories. Category-specific parameters are modelled hierarchically, allowing for partial pooling of information across categories while estimating category-specific group differences. This structure naturally accounts for multiple comparisons and quantifies the evidence for differential LLM performance across clinical information types.

For Bayesian analyses, we sample from posterior distributions using Hamiltonian Monte Carlo with 4 chains, 4000 iterations per chain after 2000 warm-up iterations. Convergence is assessed using split-*R̂<* 1.01 and bulk/tail effective sample sizes *>* 1000. All analyses reported as posterior means and 95% highest density intervals.

### Evaluation Metrics

We employ a comprehensive evaluation framework combining multiple reliability and validity measures focused on standardised SNOMED-CT codes.

#### Annotation Matching

To compare annotations between raters, we implement a two-step matching process:

1. **Primary SNOMED-CT matching**: For inter-rater reliability assessment, we first match annotations directly using their assigned SNOMED-CT codes. For each

clinical category (socio-demographics, signs, symptoms, diagnoses, treatments, risk factors, and test results), we identify matching codes between rater pairs.

1. **Secondary similarity-based matching**: For annotations without SNOMED-CT code matches, we employ the Hungarian algorithm [22] to identify optimal matching pairs. This algorithm maximises overall similarity based on weighted field comparisons: primary fields (text and context) weighted at 0.3 each, and secondary fields (qualifiers, laterality, etc.) weighted at 0.05 — 0.1 each. This differential weighting reflects the hierarchical importance of annotation components, with direct textual information carrying greater weight than supplementary qualifiers.

This hybrid approach ensures that we leverage the objective nature of SNOMED-CT codes where available, while still accommodating the need for more nuanced similarity assessments when exact code matches aren’t present.

#### Inter-rater Reliability

To assess hypotheses H1 and H2 regarding inter-rater reliability between LLMs and medical professionals, we employ Cohen’s Kappa (*κ*) as our primary reliability metric:

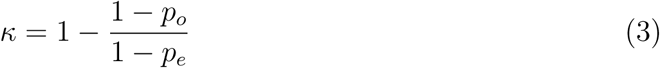

where *p_o_*represents observed relative agreement and *p_e_* expected, chance agreement.

Cohen’s Kappa accommodates multiple raters, handles missing data, and is applicable to nominal data (SNOMED-CT codes). This metric is calculated separately for Medic- Medic (M-M), Medic-LLM (M-L), and LLM-LLM (L-L) pairings to enable systematic comparison of agreement patterns necessary for testing H1 and H2. For each EHR, agreement is assessed based on the matched SNOMED-CT codes, with analyses conducted both globally (H1) and for each clinical information category (H2)(socio-demographics, signs, symptoms, diagnoses, treatments, risk factors, and test results).

#### Error Analysis

The accuracy of information extraction is evaluated through standard classification metrics, using the medical professionals’ annotations as the ground truth. For each clinical element identified, we calculate:

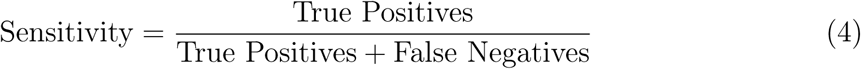

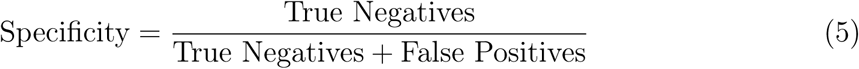

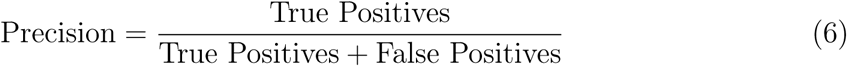

Given the critical importance of avoiding hallucinated information in clinical settings, we place particular emphasis on the False Positive Rate (FPR):

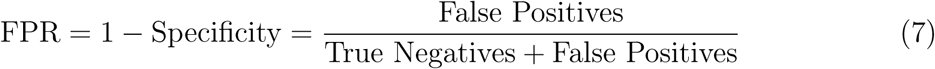

To account for multiple human annotators, we employ a permutation approach where each human annotation serves as the ground truth in turn, with results averaged across all permutations. This approach provides a robust assessment of model performance while accounting for natural variation in human annotation.

The classification metrics are calculated both globally and stratified across the seven key categories of clinical information (socio-demographics, symptoms, signs, diagnoses, treatments, risk factors, and test results). This stratification enables identification of category-specific strengths and weaknesses in model performance.

## Results

### Annotations

When considering the average number of annotations per EHR, by qualifier, the most highly annotate qualifier by humans was procedures with a mean of *µ* = 15.54 *±* 1.32 annotations per EHR, while Risk factors received the fewest annotations per EHR (*µ* = 1.01 *±* 0.23), see Figure 2a.

**Figure 2:**
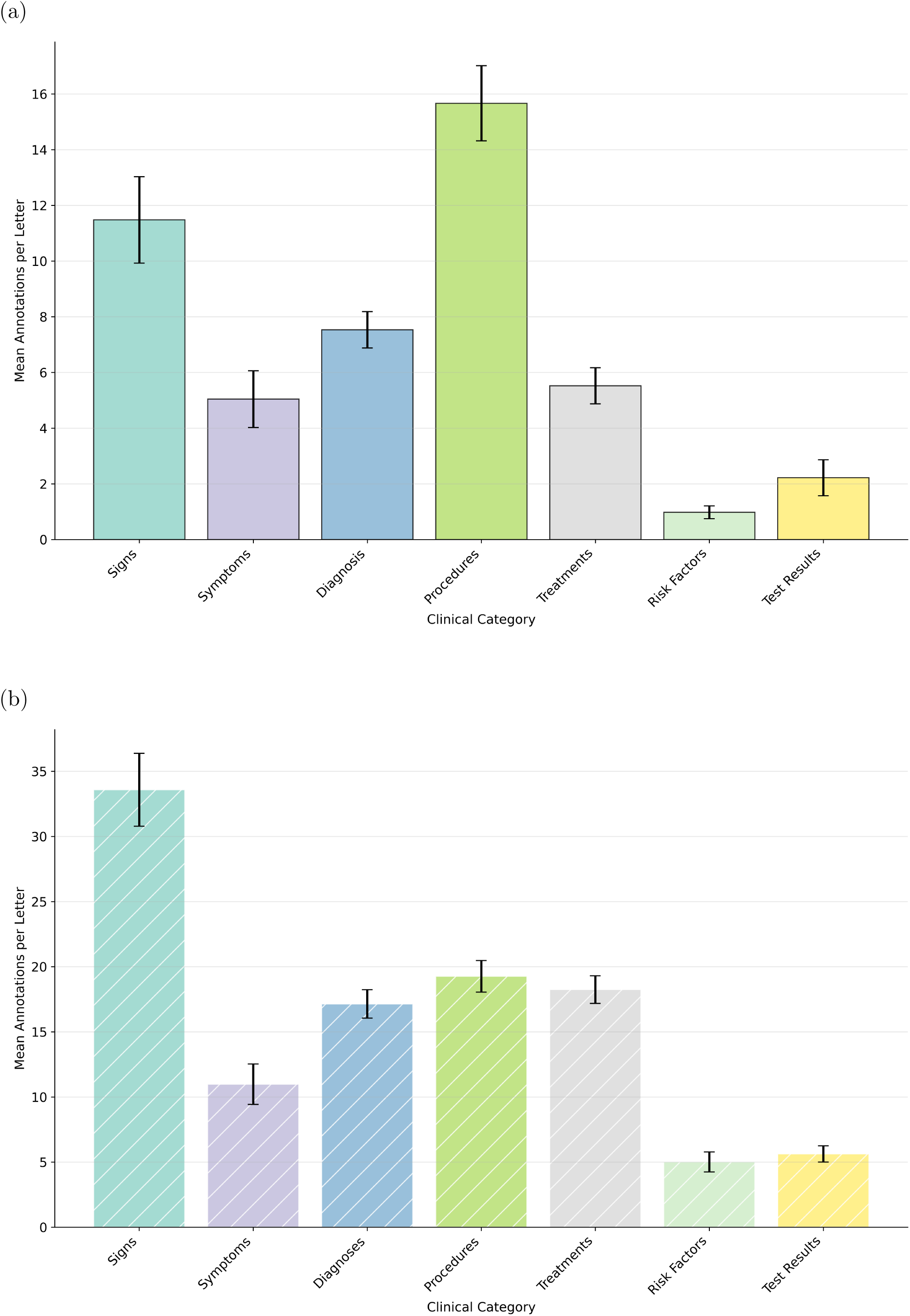
Mean number of annotations made by medical professionals (a.) and LLMs (b.) per EHR by clinical category. Error bars indicate standard error.

We tested seven LLMs on the task of annotating EHRs with clinical information and SNOMED-CT codes, analysing all 98 EHRs. Notably, LLMs consistently extracted more clinical entities per EHR than medical professionals across all categories (see figure 2b), with the most pronounced differences in risk factors (*µ* = 7.33 *±* 1.17) and signs (*µ* = 43.21 *±* 3.55).

### Inter-rater Reliability Analysis

#### Medic-Medic Comparison

Overall, when comparing all EHRs and all annotations, the average Cohen’s Kappa was *κ* = 0.752, indicating a substantial level of agreement. When considering this by qualifier, the annotations present a varied level of inter-rater reliability between *κ* = 0.429 for procedures and *κ* = 1 for test results, see Figure 3a.

**Figure 3:**
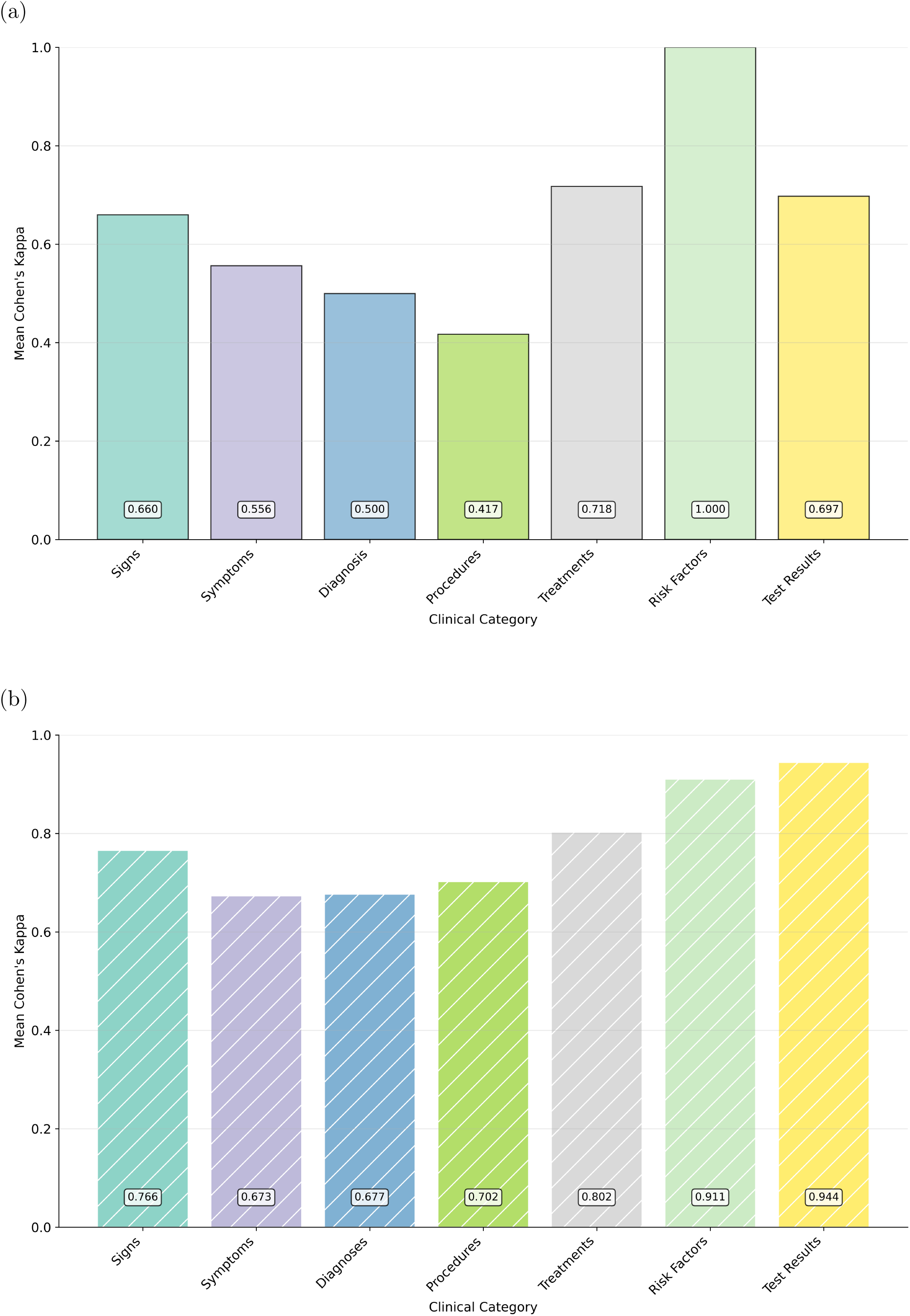
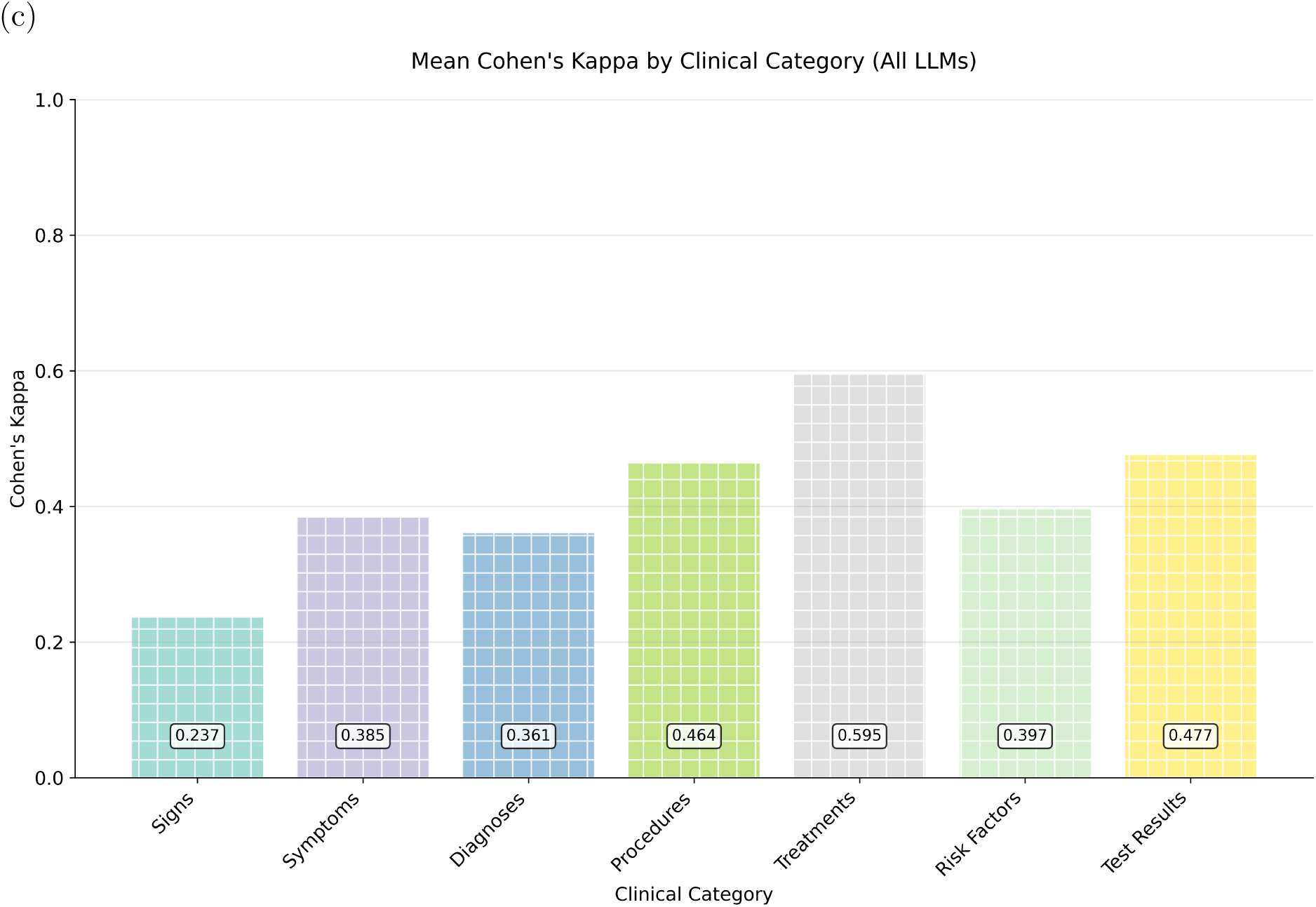
Cohen’s Kappa (*κ*) calculated for each clinical category for the human-human (a), LLM-LLM (b) and, human-LLM (c) annotation comparisons.

Most qualifiers yielded substantial agreement (Signs, Treatments and, Test Results), some highlighted moderate agreement (Symptoms and procedures) and Risk Factors yielded a perfect agreement, although that is likely due to the low occurrences of these qualifier types in the data.

#### LLM-LLM Comparison

Overall, when comparing all EHRs and all LLMs, the average Cohen’s Kappa was *κ* = 0.795, indicating a substantial level of agreement. Figure 3b shows inter-rater reliability, varying between *κ* = 0.656 for symptoms and *κ* = 0.938 for test results.

Most qualifiers yielded substantial agreement (Signs, Treatments and, Test Results), some highlighted moderate agreement (Symptoms and procedures) and Risk Factors yielded a perfect agreement.

#### Medic-LLM Comparison

Overall, when comparing all EHRs annotated by medics against all LLMs, the average Cohen’s Kappa was *κ* = 0.391, indicating a fair level of agreement. When considering this by qualifier, inter-rater reliability varied between *κ* = 0.259 for signs and *κ* = 0.500 for treatments, see Figure 3c. When examining individual model performance, substantial variation emerged across the seven LLMs tested (Table 2). Claude Sonnet 3.5 and LLAMA 3.1 8B achieved the highest overall agreement with medical professionals (*κ* = 0.529 and *κ* = 0.544, respectively), while LLAMA 3.1 405B demonstrated the lowest (*κ* = 0.260). Notably, model performance varied considerably by clinical category, with most models achieving strongest agreement for treatments (ranging from *κ* = 0.419 to *κ* = 0.778) and weakest for signs (*κ* = 0.134 to *κ* = 0.394).

**Table 2:**
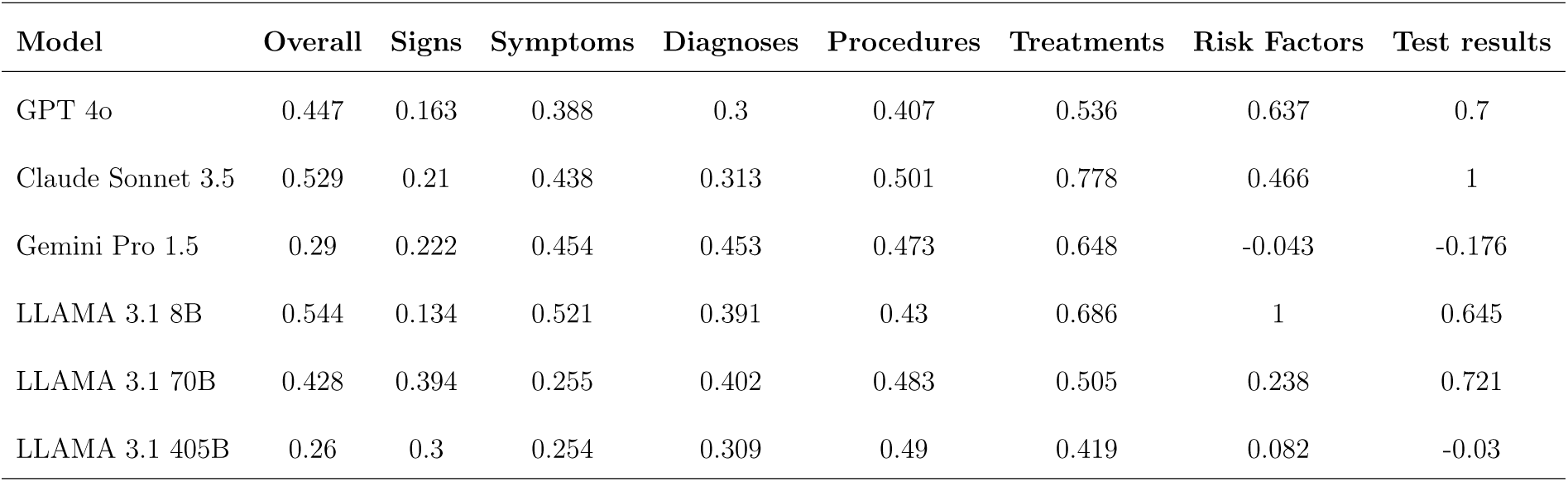
Mean Cohen’s Kappa estimates for each LLM model as compared to medical professionals’ annotations, split by clinical category.

| Model | Overall | Signs | Symptoms | Diagnoses | Procedures | Treatments | Risk Factors | Test results |
| --- | --- | --- | --- | --- | --- | --- | --- | --- |
| GPT 4o | 0.447 | 0.163 | 0.388 | 0.3 | 0.407 | 0.536 | 0.637 | 0.7 |
| Claude Sonnet 3.5 | 0.529 | 0.21 | 0.438 | 0.313 | 0.501 | 0.778 | 0.466 | 1 |
| Gemini Pro 1.5 | 0.29 | 0.222 | 0.454 | 0.453 | 0.473 | 0.648 | -0.043 | -0.176 |
| LLAMA 3.1 8B | 0.544 | 0.134 | 0.521 | 0.391 | 0.43 | 0.686 | 1 | 0.645 |
| LLAMA 3.1 70B | 0.428 | 0.394 | 0.255 | 0.402 | 0.483 | 0.505 | 0.238 | 0.721 |
| LLAMA 3.1 405B | 0.26 | 0.3 | 0.254 | 0.309 | 0.49 | 0.419 | 0.082 | -0.03 |

#### Bayesian Hypothesis Testing

To formally test our hypotheses regarding differences in inter-rater reliability between groups, we applied Bayesian hierarchical modeling to the pairwise Cohen’s Kappa values. We hypothesised that there is no significant difference in inter-rater reliability between LLMs and medical professionals when extracting SNOMED-coded clinical information from ENT EHRs. The posterior distributions revealed distinct patterns between groups (Figure 4.). Substantially lower agreement in medic-LLM pairs; the posterior mean Cohen’s Kappa for medic-medic pairs was 0.813 (95% CI: 0.755–0.860), compared to 0.659 (95% CI: 0.633–0.684) for medic-LLM pairs, mean difference of −0.154 95% CI: −0.209 to −0.091). Non-inferiority testing revealed insufficient evidence to support equivalence at any tested margin: the posterior probability that medic-LLM agreement was within 0.05 of medic-medic agreement was 0.002, increasing to only 0.045 for a margin of 0.10 and 0.430 for a margin of 0.15.

**Figure 4:**
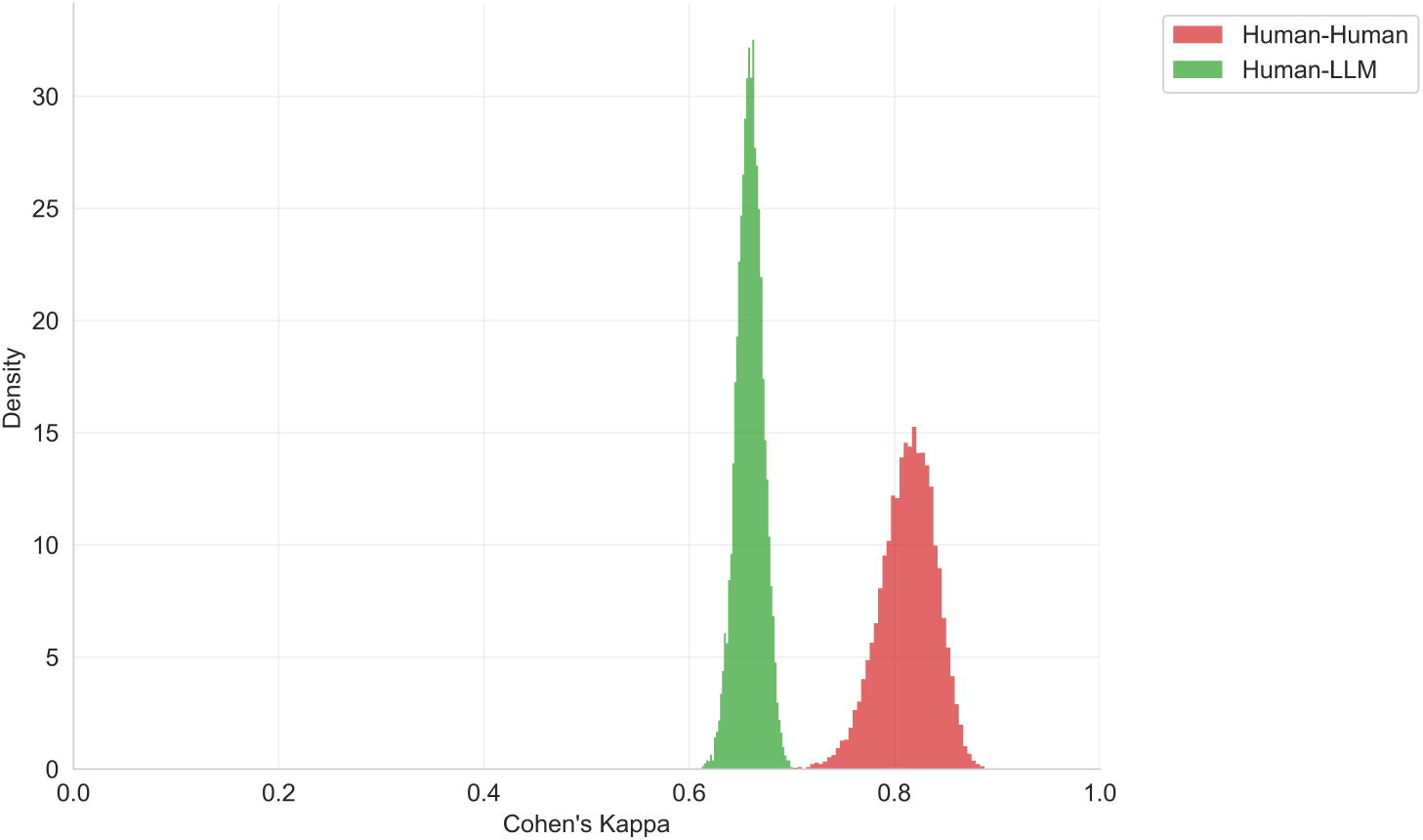
Posterior probability distributions of Cohen’s kappa estimates for inter-rater reliability. The red distribution represents Medic-Medic (M-M) inter-rater reliability with a posterior mean of 0.813 (95% CI: 0.755-0.860), while the green distribution represents Medic-LLM (M-L) inter-rater reliability with a posterior mean of 0.659 (95% CI: 0.633-0.684). Distributions were derived from Bayesian hierarchical modeling using Beta-distributed likelihood functions with weakly informative Gamma priors (*α* = 3*, β* = 1).

We hypothesised that there is no significant difference in inter-rater reliability between LLMs and medical professionals when extracting SNOMED-CT codes across different clinical information categories. The hierarchical model showed minimal group × category interaction (variance = 0.0018, 95% CI: 0.0003–0.0054), indicating consistent medic-LLM agreement deficits across categories. Medic-LLM agreement was lower than medic-medic agreement for all categories except test results, with differences ranging from −0.035 (test results) to −0.099 (diagnoses) (Figure 5).

**Figure 5:**
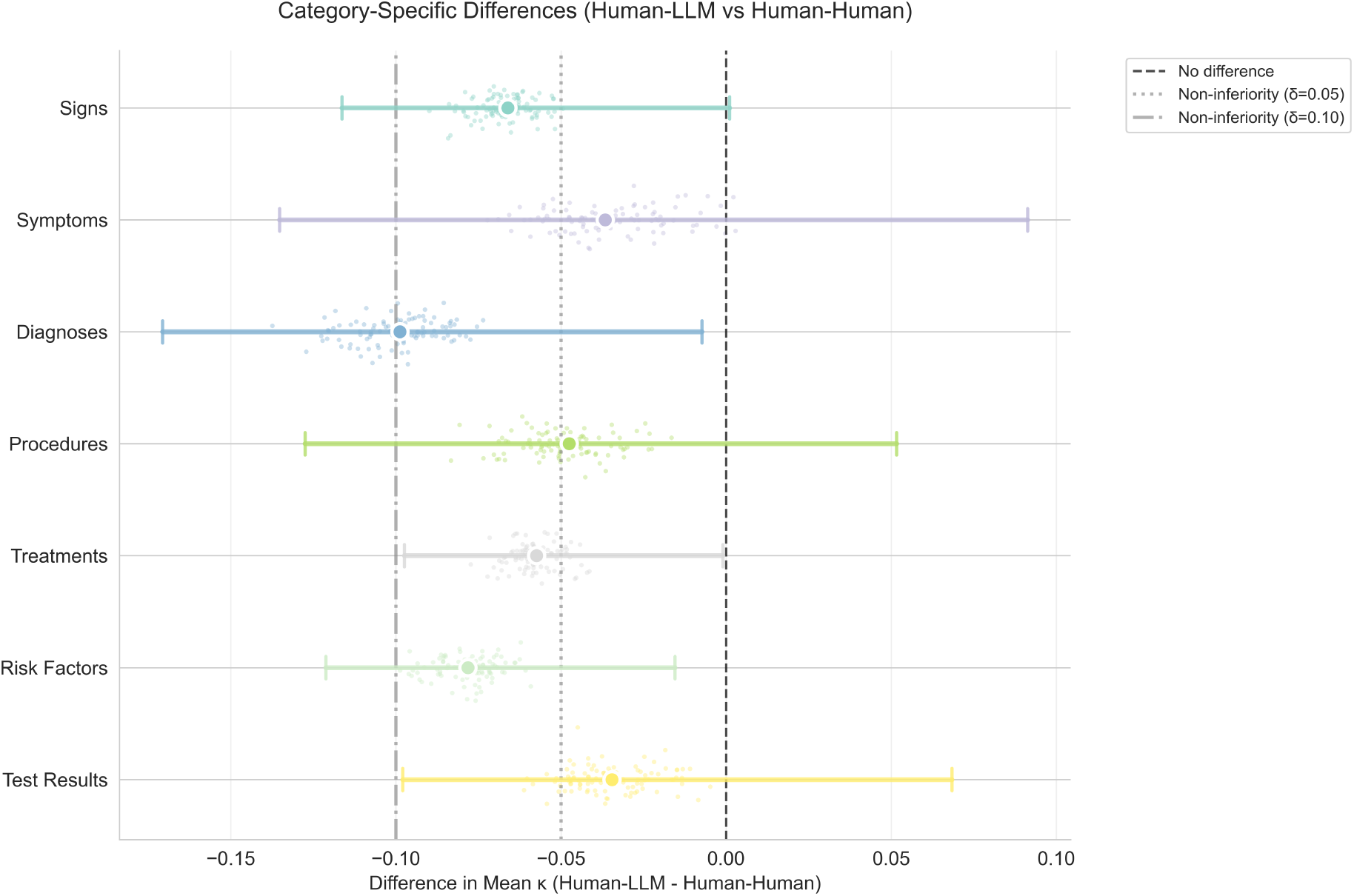
Category-specific differences in Cohen’s Kappa between human-LLM and human-human pairs. Points represent posterior means with 95% credible intervals. Vertical lines indicate non-inferiority margins at *δ* = 0.05 (dotted), 0.10 (dashed), and 0.15 (dash-dot).

Non-inferiority was achieved (>95% posterior probability) only at the 0.15 margin for most categories, and at the 0.10 margin for treatments and test results. These findings suggest that while LLM performance varies somewhat by clinical category, no category achieves parity with human-human agreement.

All models demonstrated excellent convergence (*R*^^^ *<* 1.01, ESS > 10,000) across all parameters, providing confidence in the posterior estimates.

### Error Analysis and Classification Performance

To evaluate the accuracy of LLM information extraction, we analysed classification performance across all clinical categories for a chosen model (GPT-4o). Manually assessing all annotations to establish a ground truth, we assessed model performance through standard classification metrics with particular attention to false positive rates given their clinical significance.

The LLMs demonstrated robust performance in extracting clinical information from ENT, hearing and balance EHRs, achieving an overall accuracy of 86.9% (95% CI: 86.3% – 87.4%) across 13,656 evaluated annotations (Figure 6.). The models exhibited particularly high precision (97.0%, 95% CI: 96.6%–97.3%), indicating strong reliability when identifying clinical elements. However, recall was moderately lower at 84.9% (95% CI: 84.2%–85.6%), suggesting that while extracted information was highly accurate, some clinical elements were missed. The specificity of 92.6% (95% CI: 91.6%–93.3%) and corresponding false positive rate of 7.5% demonstrate acceptable but not negligible rates of hallucinated content.

**Figure 6:**
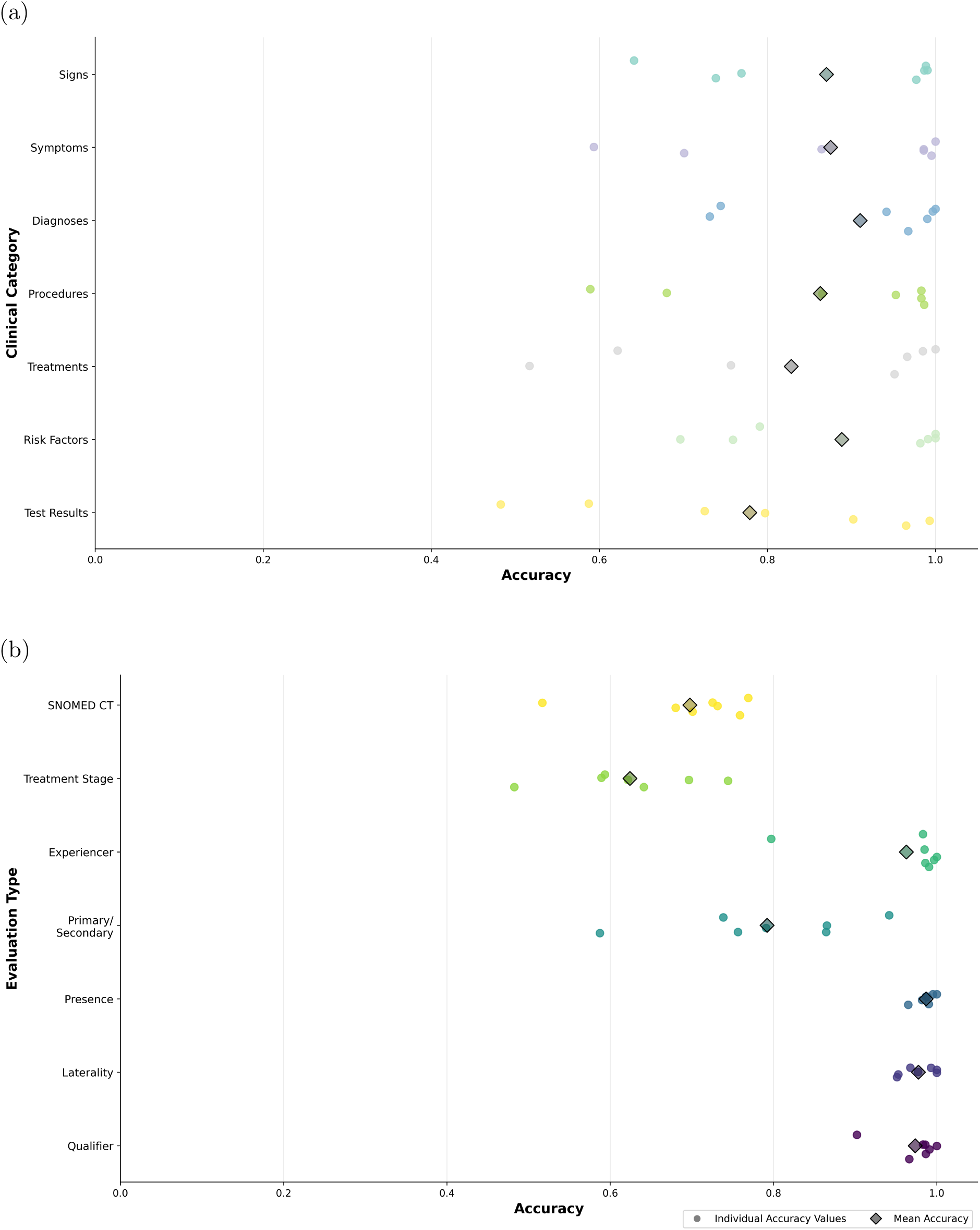
Accuracy of GPT-4o’s annotations across ENT EHRs and notes split by clinical category (a) and information type (b).

#### Performance by Clinical Category

Classification performance varied substantially across clinical information categories (Figure 6a). Diagnoses demonstrated strong performance with 91.0% accuracy (95% CI: 89.7% – 92.2%) and balanced precision-recall characteristics (precision: 97.7%, recall: 91.4%). Similarly, symptoms and signs showed consistent extraction quality, with accuracies of 87.5% and 87.0% respectively, though signs exhibited a slightly higher false negative rate (16.1% vs 14.3% for symptoms).

More challenging categories included treatments (82.8% accuracy, 95% CI: 81.2% – 84.5%) and test results (77.9% accuracy, 95% CI: 75.6% – 80.4%). The lower performance in test results extraction was primarily driven by reduced recall (70.4%), indicating difficulty in identifying all relevant test information within the clinical narratives. Notably, test results showed the highest false negative rate (29.6%) among all categories, suggesting that LLMs may struggle with the varied formats and contextual embedding of laboratory and diagnostic findings in clinical text.

Risk factors, despite being the least frequently annotated category, maintained reasonable performance (88.9% accuracy) with exceptionally high precision (99.0%, 95% CI: 97.9%–99.8%). The low false positive rate (1.0%) for risk factors indicates that LLMs rarely hallucinate these clinical elements, though the false negative rate of 17.3% suggests some relevant risk factors remain unidentified. Performance also varied when stratified by evaluation type, with accuracy ranging from 77% to 89% across different validation frameworks (Figure 6b). This variation reflects the different methodological approaches used to assess annotation quality, with some evaluation types showing more conservative performance estimates than others. Complete performance metrics for all clinical categories and evaluation types are provided in Supplementary Material 1.

## Discussion

This study provides the first systematic evaluation of LLM performance in clinical information extraction using standardised inter-rater reliability metrics, with ENT EHRs serving as a test case for broader medical applications. Our findings demonstrate that while LLMs achieve substantial agreement with medical professionals, they fall short of human-human reliability levels (*κ* = 0.659 vs *κ* = 0.813), with important variations across clinical information categories. These results have significant implications for AI deployment in clinical documentation workflows across medical specialties.

### Principal Findings and Their Significance

Our Bayesian hierarchical analysis conclusively rejected the hypothesis of equivalent performance between LLMs and medical professionals, with only a 4.5% probability that human-LLM agreement falls within 0.1 of human-human agreement. While this performance gap was consistent across clinical categories, the pattern of differences provides important insights. LLMs performed best for treatments and test results, typically explicit and standardised in clinical documentation, but showed larger gaps for signs and diagnoses that require nuanced clinical interpretation.

The finding that LLMs consistently extracted more clinical entities than medical professionals, particularly for risk factors (7.33 vs 1.01 annotations per EHR), reveals a fundamental difference in information processing. This aligns with previous observations that physicians often de-prioritize primary prevention and risk factor documentation in favor of addressing acute conditions and disease management [24]. This divergence is particularly relevant as medicine evolves toward precision therapeutics requiring comprehensive phenotyping [14, 16]. LLMs may capture information that clinicians consider implicit or contextually less relevant, suggesting they could complement human expertise by ensuring comprehensive data capture for phenotyping algorithms and clinical decision support systems.

### Implications for Clinical Implementation

The high precision (97.0%) but moderate recall (84.9%) observed across LLMs suggests a clear implementation strategy: LLMs are best suited for augmentative roles where accuracy is paramount, with human oversight ensuring completeness. This aligns with emerging collaborative human-AI workflows where LLMs handle initial extraction and clinicians focus on verification and contextualisation [25, 26]. The 7.5% false positive rate, while low, remains non-trivial for clinical applications and reinforces the need for human oversight.

Our finding that different LLMs share similar biases (LLM-LLM *κ* = 0.795) has important implications for healthcare systems considering AI adoption. This consistency suggests that performance limitations reflect fundamental challenges in clinical language understanding rather than model-specific issues, indicating that prompt engineering or model selection alone may not overcome these limitations without architectural innovations or domain-specific training [27, 28].

### Study Limitations and Generalisability

Several limitations merit consideration. The most significant limitation concerns our use of MTSamples as the data source. While these publicly available documents enable reproducible research, they may not fully represent the heterogeneity of modern EHR documentation across different institutions and systems. MTSamples documents were created for educational purposes and may differ from contemporary clinical documentation in structure, completeness, and complexity.

Our medical annotators, while all qualified doctors from various hospitals and backgrounds, may not capture the full spectrum of clinical documentation practices across different healthcare settings, experience levels, and geographic regions. The annotation process itself, conducted in a research context, may differ from real-world clinical documentation workflows where time pressures and competing priorities influence information extraction.

The static nature of our evaluation does not capture the potential for improvement through fine-tuning or few-shot learning [29]. Future work should investigate whether domain-specific adaptation can narrow the performance gap, particularly for categories showing the largest discrepancies.

Despite these limitations, the consistent patterns observed across multiple LLM architectures and the systematic nature of our evaluation provide valuable insights into current LLM capabilities for clinical information extraction.

### Future Directions and Broader Impact

This work establishes a rigorous framework for evaluating clinical AI systems that can accelerate responsible AI deployment in healthcare. The performance gap identified highlights opportunities for advancing AI capabilities through incorporating clinical knowledge graphs [30], developing specialty-specific models, or designing architectures that better model hierarchical medical reasoning.

As healthcare systems increasingly adopt AI technologies[31], our findings provide evidence based guidance for implementation. The category-specific performance profiles can inform targeted deployment strategies, prioritising AI assistance where performance is strongest while maintaining human oversight for complex clinical reasoning tasks. This balanced approach ensures that AI enhances rather than replaces human clinical expertise, supporting the evolution toward more comprehensive, data-driven healthcare while maintaining the nuanced judgment that defines excellent clinical practice [32].

## Declarations

### Ethics approval and consent to participate

This study analysed publicly available, de-identified clinical documentation from MTSamples. The annotation and data extraction tasks were performed by the authors who are medical professionals, all of whom provided informed consent for their participation in this research. Following consultation with the University College London Research Ethics Committee guidelines, it was determined that formal ethics approval was not required as this study design is analogous to systematic review methodology where authors serve as data extractors analysing publicly available data. The study adheres to the principles of the Declaration of Helsinki for research involving human data.

While ethics approval was not required, we registered our data processing protocols with University College London’s Data Protection Office to ensure appropriate data handling practices. This registration was approved under Reference: Z6364106/2025/01/13, confirming our compliance with data protection regulations and best practices for secondary data processing.

### Consent for publication

Not applicable.

### Authors’ contributions

L.B. was responsible for conceptualisation, resources, software, data curation, formal analysis, validation, investigation, visualisation, methodology, project administration, writing - original draft and writing - review & editing. N.J. contributed to conceptualisation, data curation, annotation and writing - review & editing. A.N., L.D., E.M., T.R., R.P., K.P., I.M., L.F.T., S.S., K.M., E.B., C.D., and H.F. performed clinical annotation and data curation. A.P. and Z.K. were responsible for data curation. K.C. handled data curation, validation, and formal analysis. A.S. provided supervision, funding acquisition and writing - review & editing. N.M. contributed to conceptualisation, methodology, investigation, supervision, and writing - review & editing. All authors read and approved the final manuscript.

### Funding

This study was funded by NIHR BRC UCLH (IS-BRC-1215-20016) Hearing Health theme. The funder played no role in study design, data collection, analysis and interpretation of data, or the writing of this manuscript.

### Competing interests

K.M. is employed by Ufonia Ltd.

All other authors declare no financial or non-financial competing interests.

### Data availability

All data associated with the EHRs, human and large language model annotations and SNOMED codes are available publicly at https://github.com/evidENT-AI/LLM-EHR-IRR.git.

### Code availability

All code to undertake the data extraction, large language model pipeline and resultant data analysis is available publicly at https://github.com/evidENT-AI/LLM-EHR-IRR.git.

## Notes

### Competing Interest Statement

The authors have declared no competing interest.

### Author Declarations

The study used ONLY openly available human data that were originally located at: https://www.mtsamples.com/site/pages/browse.asp?type=100-ENT%20-%20Otolaryngology

